# Ethnic differences in the incidence of clinically diagnosed influenza: an England population-based cohort study 2008-2018

**DOI:** 10.1101/2021.01.15.21249388

**Authors:** Jennifer A. Davidson, Amitava Banerjee, Rohini Mathur, Mary Ramsay, Liam Smeeth, Jemma Walker, Helen McDonald, Charlotte Warren-Gash

**Author notes:** **Corresponding author details** Name: Jennifer Davidson. Authors contributed equally (Helen McDonald, Charlotte Warren-Gash).

## Abstract

We used primary and linked secondary healthcare data to investigate the incidence of clinically diagnosed influenza/influenza-like-illness (ILI) by ethnicity in England from 2008-2018. We identified higher incidence rate ratios for influenza/ILI among South Asian (1.70, 95% CI 1.66-1.75), Black (1.48, 1.44-1.53) and Mixed (1.22, 1.15-1.30) groups compared to White ethnicity.

People from ethnic minority backgrounds are represented disproportionately among patients with severe COVID-19. Recent research has found people of Black and South Asian ethnicity have increased risk of SARS-CoV-2 infection, COVID-19-related hospitalization and death, independent of deprivation, occupation, household size, and underlying health conditions(1,2).

The COVID-19 pandemic has reinforced the importance of seasonal influenza vaccination. By preventing influenza-related hospitalization, vaccination can minimize the risk of hospital-acquired COVID-19 (co-) infection for these individuals and reduce health service pressures, particularly the need for isolation of patients with respiratory symptoms awaiting COVID-19 test results.

In the UK, influenza vaccine is routinely recommended for adults aged ≥65 years, or people <65 years with underlying health conditions. These recommendations formed the basis of the original guidance to identify patients at moderate- and high-risk of COVID-19. Influenza vaccine recommendations were expanded for the 2020/21 season to include all adults ≥50 years(3). However, vaccine uptake among clinical risk groups is low, particularly for Black and Mixed Black ethnic groups(4). In addition, people of non-White ethnicity have higher risk of severe outcomes following influenza infection(5,6). It is unclear whether this is driven by the risk of infection or complications, with most research focused on distal outcomes rather than initial infection risk.

Here we investigate the incidence of influenza and influenza-like-illness (ILI) by ethnicity from 2008-2018 among people not eligible for routine influenza vaccination, to consider disparities in infection risk.

## METHODS

We conducted a retrospective cohort study using anonymized primary care data from the UK Clinical Practice Research Datalink (CPRD) GOLD and Aurum databases(7,8) with linked secondary care data from the Hospital Episodes Statistics Admitted Patient Care (HES APC) database and death data from the Office for National Statistics. The data include diagnoses, prescriptions, immunizations and demographics.

We included adults aged 40-64 years registered at a CPRD contributing practice between 01/09/2008 and 31/08/2018. We excluded patients with a health condition indicative of influenza vaccination eligibility (Supplementary Table 1), and those who had ever received pneumococcal vaccination, or influenza vaccination in the 12 months before baseline. Follow-up started at the latest of 12 months after current registration, up-to-research-standard (GOLD only), 40^th^ birthday, or 01/09/2008. Follow-up ended at the earliest of; a new diagnosis of a condition conferring eligibility for vaccination, pneumococcal or influenza vaccination, death, transfer out, the practice’s last data collection, 65th birthday, or 31/08/2018.

Self-reported ethnicity was captured in CPRD and supplemented with HES APC if missing in CPRD. We grouped ethnicity into the five and 16 census categories of White (British, Irish, Other White), South Asian (Indian, Pakistani, Bangladeshi, other Asian), Black (African, Caribbean, other Black), Other (Chinese, all other), and Mixed (White and Asian, White and African, White and Caribbean, Other Mixed).

Influenza/ILI infection was identified from diagnostic codes in CPRD and HES APC. In a second analysis, we expanded our outcome definition to acute respiratory infection (ARI), additionally including codes for pneumonia, acute bronchitis, or other acute infections suggestive of lower respiratory tract involvement (all codes listed here, DOI: *awaiting*).

We calculated crude annual infection incidence rates by ethnic group with age- and sex-stratification. Multivariable Poisson regression models with random effects, to account for multiple infections in the same patient, were used to estimate any ethnic disparities in infection risk. Our main analysis adjusted for age, sex, and influenza season/year. A second model additionally adjusted for region of residence and socioeconomic status (based on patient-level Townsend score quintiles), which may both confound and mediate an association between ethnicity and infection.

## RESULTS

Our cohort included 3,735,308 patients (Supplementary Figure 1), of whom 87.6% were White (n=3,271,115), 5.2% South Asian (n=196,262), 4.2% Black (n=157,075), 1.9% Other (n=69,440), and 1.1% Mixed (n=41,416) (Supplementary Table 2). We excluded 511,682 (12.0%) patients with no recorded ethnicity; this group had longer follow-up, fewer consultations and were more likely to be male than the included study population (Supplementary Table 2). 16-category ethnicity was known for 3,035,689 of the cohort (with HES ethnicity breakdown beyond white and mixed not available), of whom 76.3% were White British, 0.9% Irish, 7.7% Other White, 2.6% Indian, 1.3% Pakistani, 0.4% Bangladeshi, 2.2% Other Asian, 2.8% African, 1.5% Caribbean, 0.9% Other Black, 0.7% Chinese, and 1.5% Other (Supplementary Table 3). Non-White populations were younger and resided in more deprived areas than the White population, while a higher proportion of the White population were obese.

**Figure 1.**
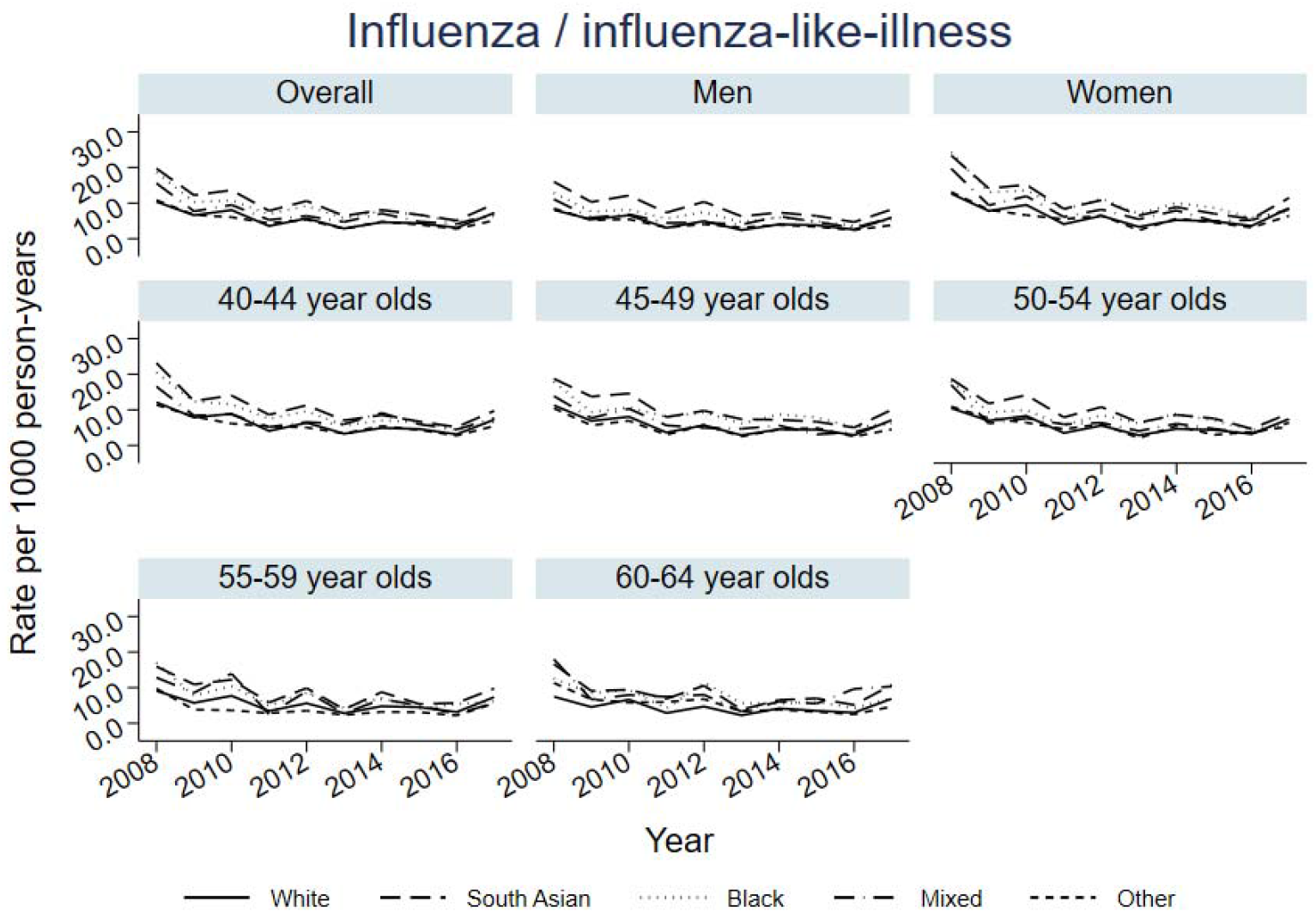
Overall, age- and sex-stratified annual incidence rates for influenza / influenza-like-illness by ethnic group

We identified 102,316 influenza/ILI episodes recorded among 94,623 patients, and 560,860 ARI episodes among 421,349 patients. The rate of influenza/ILI was highest in the South Asian group (9.6 per 1,000 person-years) followed by the Black group (8.4 per 1,000 person-years) (Figure 1, Supplementary Table 4). In all ethnic groups the influenza/ILI rates were higher in women than men and decreased with age.

After adjustment for age, sex and year, the incidence rate ratio (IRR) for influenza/ILI was higher for South Asian (1.70, 95% CI 1.66-1.75), Black (1.48, 95% CI 1.44-1.53), and Mixed (1.22, 95% CI 1.15-1.30) groups compared to the White group (Figure 2, Supplementary Table 4). When broken down into the 16 categories, the IRR for influenza/ILI was higher in all groups included in the South Asian, Black and Mixed broad ethnic classifications, with the highest IRR in the Bangladeshi group (2.26, 95% CI 2.05-2.49). After additional adjustment for deprivation and region, results remained similar.

**Figure 2.**
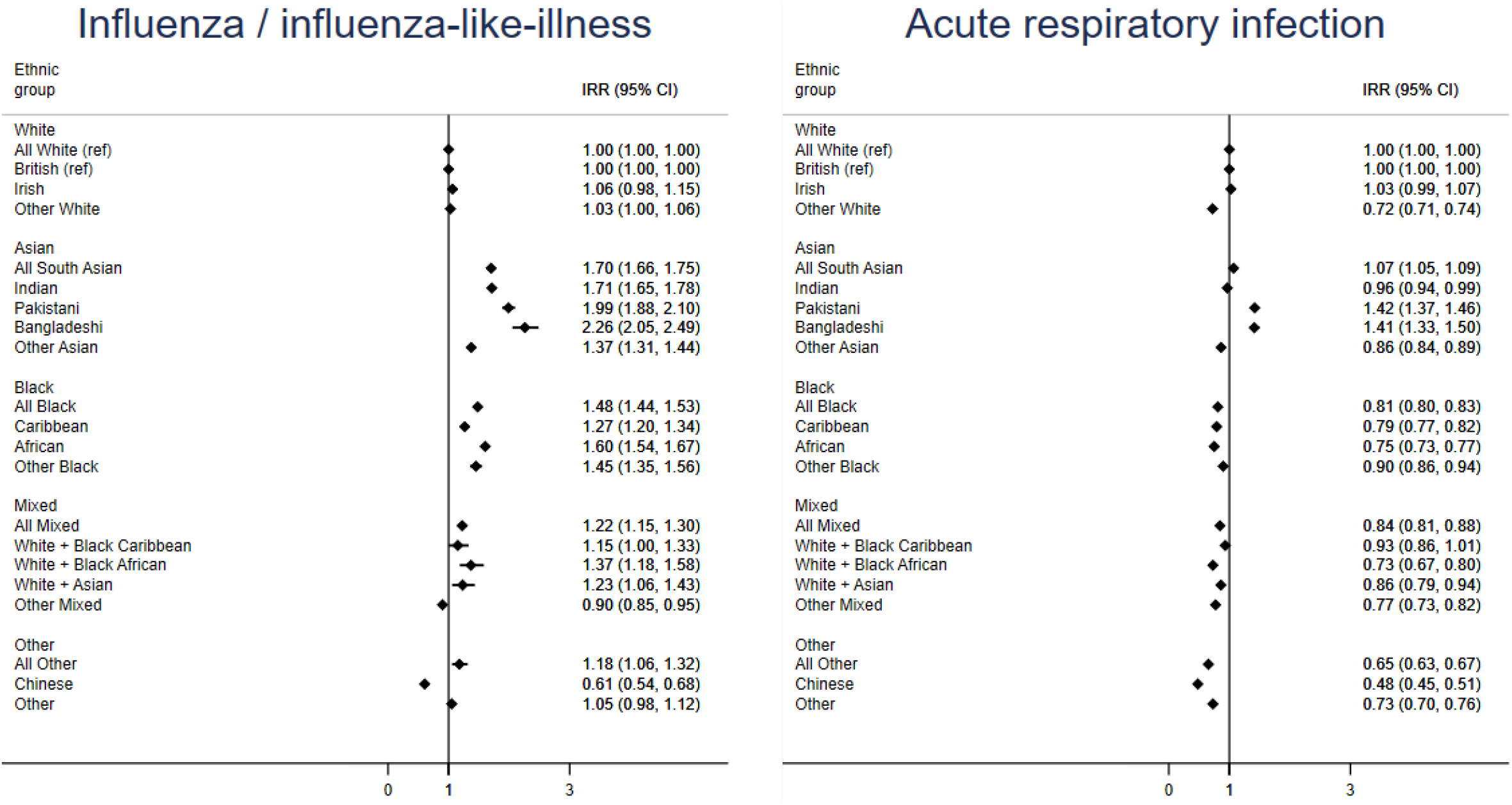
Ethnic differences in the incidence ratio risks of influenza / influenza-like-illness and acute respiratory infections All White is the reference category for comparison of ethnicity in 5 categories. British is the reference category for comparison of ethnicity in 16 categories. Models were adjusted for 5-year age band, sex, year.

For ARI, the IRR was higher in the South Asian group (1.07, 95% CI 1.05-1.09) when compared to the White group, but lower in the Black (0.81, 95% CI 0.80-0.83), Mixed (0.84, 95% CI 0.81-0.88) and Other (0.65, 95% CI 0.63-0.67) groups. Using the 16 categories, the IRR for ARI was only higher for the Pakistani (1.42, 95% CI 1.37-1.46) and Bangladeshi (1.41, 95% CI 1.33-1.50) groups when compared with the White British group.

## DISCUSSION

We showed an increased rate of influenza/ILI among Black, South Asian and Mixed groups. Specifically, those of Indian, Pakistani, Bangladeshi and African ethnicity had the highest rate compared to the White British group. When using our broader outcome of ARI, we only found an increased rate in the South Asian group with decreased rates in Black, Mixed and Other groups.

Our results suggest influenza infection risk differs between White and non-White groups. Such findings are consistent with studies of other acute viral respiratory infections including those which investigated the ethnic disparities in severe influenza outcomes, particularly during the 2009 H1N1 pandemic(5,6) as well as studies of COVID-19 infection risk and severe outcomes(1).

Our study was conducted among patients not eligible for vaccination, and so disparities cannot be explained by differences in vaccine uptake or effectiveness: there are potentially even larger ethnic differences in influenza incidence among those eligible for influenza vaccine due to inequalities in chronic disease patterns. Since social mixing and household contact are important considerations for influenza/ILI transmission our findings are relevant to the whole population. People of non-white ethnicity tend to live in larger, multi-generational households with extended kinship and social networks(9,10). Therefore, understanding ethnic disparities in respiratory infections across both high- and low-risk populations remains important for preventing hospitalizations.

Here we have presented results of a large population-based cohort study using nationally representative data. Excluding patients eligible for influenza vaccination due to chronic medical conditions should have reduced confounding. Nevertheless, our study may be impacted by some limitations. Under-diagnosis of health conditions may differ by ethnicity, with people from some ethnic groups less likely to be excluded from our study population but more likely to have an undiagnosed, and therefore unmanaged condition, which may affect influenza risk. Ethnicity may be less well recorded in GP records for individuals without a chronic condition requiring frequent consultation, but financial incentivization between 2006-2011 boosted completion in GP records. Using hospital data boosted the completeness of ethnicity recording in our study population from 74% to 88%. Influenza infection in our study was based on clinical diagnosis. Clinically identified influenza depends not only on healthcare attendance but also clinical coding practices, both of which may be associated with ethnicity. However, our results are consistent with other studies which used laboratory-confirmed measures of acute viral respiratory infections(1,5,6). Our differing results for influenza/ILI and ARI outcomes may be attributable to the lack of specificity of ARI codes for influenza. We excluded individuals with known risk factors for influenza, it may be that other conditions are relevant risk factor for ARI generally.

Ethnic inequalities in the incidence of respiratory infections could arise because of differences in risk of exposure, driven by factors such as occupation and household composition(11), as well as inequalities in access to care. We excluded children who are a key driver for influenza transmission; examining ethnic inequalities for infection risk in children is an area for future research. Unequal access to treatments will also affect the likelihood of adverse outcomes after infection.

The COVID-19 pandemic has highlighted ethnic inequalities in infection risk, which our study found are also present for influenza. This reinforces the urgency of addressing lower influenza vaccine uptake among minority ethnic groups, which could be combined with public health interventions to promote equal uptake of COVID-19 vaccination(12).

## Author contributions

H.M. and M.R. conceived the study idea. J.A.D. lead the design of the study with contributions from R.M., J.W., H.M. and C.W.-G. J.A.D. conducted the analysis. J.A.D., H.M. and C.W.-G. wrote the original manuscript draft. All authors reviewed and commented on the manuscript, and approved the final version.

## Data Availability

No additional unpublished data are available as this study used existing data from the Clinical Practice Research Datalink (CPRD) database that is only accessible to researchers with protocols approved by the CPRD's Independent Scientific Advisory Committee.

## Acknowledgements

This study is based in part on data from the Clinical Practice Research Datalink obtained under licence from the UK Medicines and Healthcare products Regulatory Agency. The data is provided by patients and collected by the NHS as part of their care and support. The interpretation and conclusions contained in this study are those of the authors alone. The study was approved by the Independent Scientific Advisory Committee (Protocol number: 19_209A2).

We would like to thank Vanessa Saliba, Consultant Epidemiologist in the department of Immunisation and Countermeasures at Public Health England, for her review of our study protocol and manuscript.

## Ethical approval

The study was approved by the London School of Hygiene and Tropical Medicine Research Ethics Committee (Reference: 17894) and by the CPRD Independent Scientific Advisory Committee (Protocol number: 19_209A2).

