## Supplementary Appendix for "Ethnic differences in the incidence of clinically diagnosed influenza: an England population-based cohort study 2008-2018"

### Supplementary Table 1. Definitions used for developing exclusion conditions using CPRD code lists*

| Health condition | Study definition |
| --- | --- |
| Cardiovascular disease (CVD) | Any previous clinical diagnosis, major intervention for, or clinical review specific to CVD including heart disease (congenital or otherwise), heart failure, stroke or transient ischaemic attack. |
| Chronic liver disease | Any previous clinical diagnosis of, or clinical review specific to, chronic liver disease including cirrhosis, oesophageal varices, biliary atresia and chronic hepatitis. |
| Chronic kidney disease (CKD) | Any previous clinical diagnosis of, or clinical review specific to, CKD stages 3-5, history of dialysis or renal transplant in Gold or Aurum. Or with estimated glomerular filtration rate (eGFR) to classify CKD stage 3-5 in Gold. |
| Chronic respiratory disease | Any previous clinical diagnosis of, or clinical review specific to, chronic respiratory disease, including chronic obstructive pulmonary disease, emphysema, bronchitis, cystic fibrosis, or fibrosing interstitial lung diseases. |
| Asthma | Any previous clinical diagnosis of, or clinical review specific to, asthma with at least two prescriptions of inhaled steroids in the year before baseline. Or any previous hospitalisation for asthma. |
| Chronic neurological disease | Any previous clinical diagnosis of, or clinical review specific to, a neurological disease such as Parkinson’s disease, motor neurone disease, multiple sclerosis (MS), cerebral palsy, dementia or a learning/intellectual disability. |
| Diabetes mellitus | Any previous diagnosis of, or clinical review specific to, diabetes mellitus, or with a prescription for medication used to treat diabetes. |
| Asplenia/sickle cell disease | Any previous clinical diagnosis of, or clinical review specific to, asplenia or dysfunction of the spleen (including sickle cell disease but not sickle cell trait). |
| Severe obesity | Latest body mass index before baseline was ≥40 kg/m^2^. |
| Immunosuppression | Any previous clinical diagnosis of, or clinical review specific to, HIV, solid organ transplant or other permanent immunosuppression (such as genetic conditions compromising immune function). |
|  | In the two years before baseline: clinical diagnosis of, or clinical review specific to, aplastic anaemia or haematological malignancy, or receiving a bone marrow or stem cell transplant. |
|  | In the year before baseline: previous clinical diagnosis of, or clinical review specific to, other/unspecified immune deficiency or receiving chemotherapy or radiotherapy. |
|  | In the year before baseline: prescription of biological therapy or at least 2 prescriptions for oral steroids or other immunosuppressants including DMARDS, Methotrexate, Azathioprine, or corticosteroid injections. |

* all codes listed here, DOI: *awaiting*

### Supplementary Table 2. Baseline characteristics by five category ethnic group

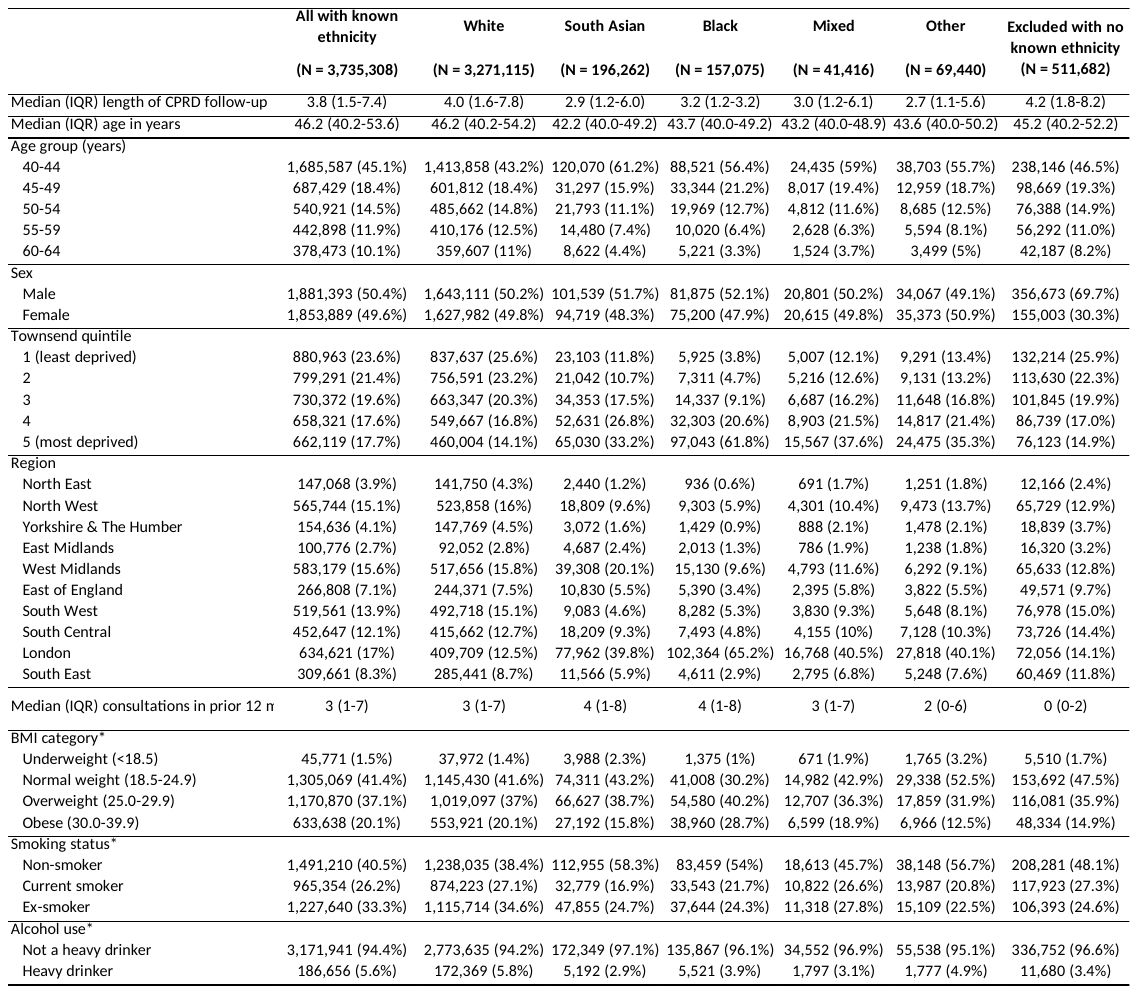

*Closest measure before baseline

### Supplementary Table 3. Baseline characteristics by 16 category ethnic group

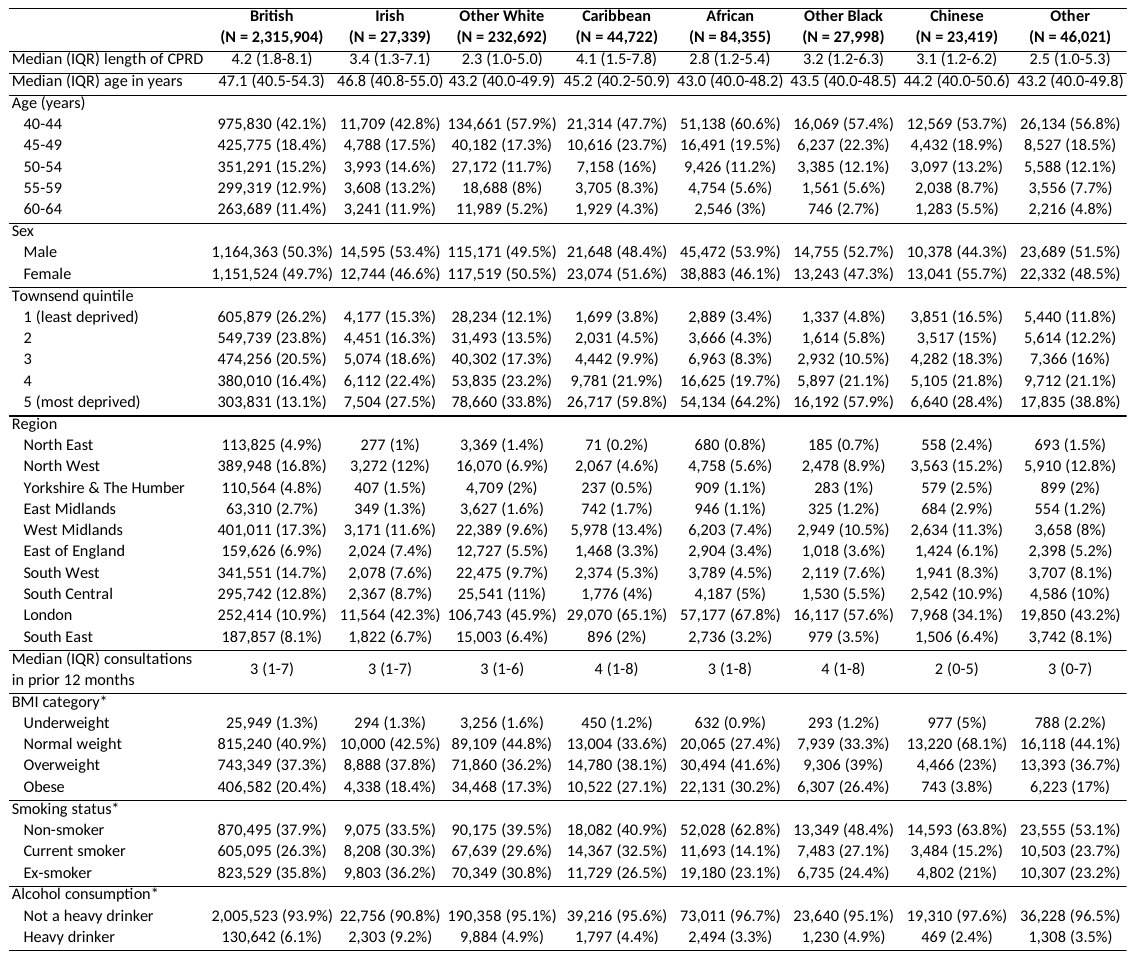

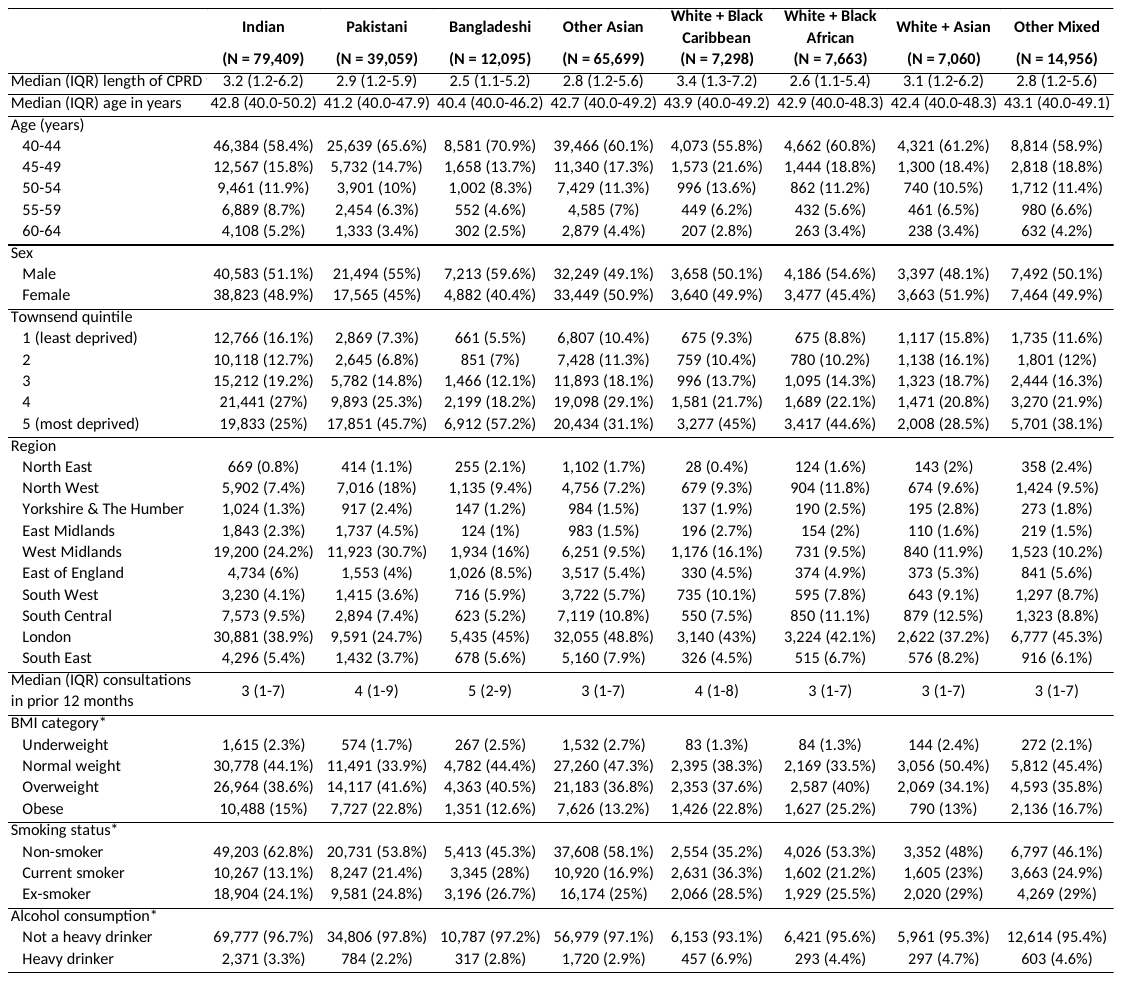

*Closest measure before baseline

Supplementary Table 4. Incidence rate ratios for influenza / influenza-like-illness and acute respiratory infections

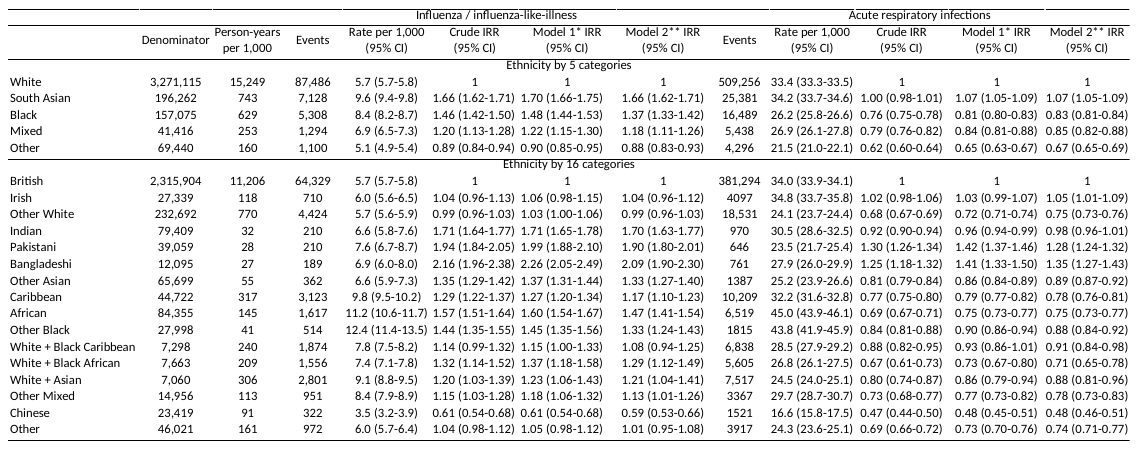

*Model 1 is adjusted for 5-year age band, sex and year. All LRT p-values <0.001

**Model 2 is adjusted for 5-year age band, sex, year, Townsend deprivation quintile and region of residence. All LRT p-values <0.001

Supplementary Figure 1. Study population flow chart

**Unknown ethnicity**

n=511,682

**Final study population**

**n=3,735,308**

**Further patient exclusion criteria**

- Removed from Gold dataset if practice also in Aurum dataset (n=644,395)
- Existing vaccine eligible condition (n=971,788)
- Previous influenza or pneumococcal vaccination (n=211,114)
- ONS death date before study follow up starts (n=1,034)

**Study population after exclusions**

**n=4,246,990**

**Met patient inclusion criteria**

- With HES linked data
- Aged 40-64 years 01/09/2008-31/08/2018
- In CPRD follow-up 01/09/2008-31/08/2018

**n=6,075,321**
